# Multi-population genome-wide association meta-analysis of acute and chronic pancreatitis

**DOI:** 10.64898/2026.09.16.26363222

**Authors:** Samuel Khodursky, Renae Judy, Sarah Abramowitz, Michael G. Levin, Scott M. Damrauer

**Affiliations:** Department of Surgery, Perelman School of Medicine at the University of Pennsylvania, Philadelphia, PA, USA; Corporal Michael J. Crescenz VA Medical Center, Philadelphia, PA, USA; Division of Cardiovascular Medicine, Department of Medicine, University of Pennsylvania, Perelman School of Medicine, Philadelphia, PA, USA; Department of Genetics, Perelman School of Medicine at the University of Pennsylvania, Philadelphia, PA, USA

## Abstract

Acute (AP) and chronic (CP) pancreatitis are major causes of gastrointestinal morbidity. However, the genetic architectures of AP and CP remain incompletely defined, as does the degree to which the two conditions share a common genetic basis. Here we report a multi-population GWAS meta-analysis of both AP and CP across five biobanks, comprising 23,292 individuals with and 1,748,420 individuals without AP across five populations and 9,866 individuals with and 1,627,720 individuals without CP across three populations. We identified 16 and 14 genome-wide significant loci for AP and CP, respectively. Combining nearest-gene assignment, fine-mapping, MAGMA, and colocalization with pancreatic eQTLs, we prioritized 7 genes for AP and 5 genes for CP using two or more approaches, including three previously unreported candidates: *TM4SF4* and *TCIM* for AP, and *FFAR4* for CP. We demonstrated that AP and CP are strongly correlated at the genome-wide level (*r_g_*=0.89); however, we identified 44 regions with significantly lower correlations, including a region on chromosome 18 containing *BCL2* with a negative local correlation. Plasma proteome-wide Mendelian randomization (MR) found 15 proteins whose genetically predicted plasma levels were associated with AP and 6 associated with CP, including ABO, whose measured levels likely proxy non-O blood type. Consistent with this, we found that non-O blood type was modestly but significantly associated with both AP and CP in an analysis of electronic health records from over 650,000 individuals. Finally, drug-target MR identified 38 and 45 target genes whose genetically predicted expression levels were associated with genetic liability for AP and CP, respectively. Among these, *DRD2* and *BCHE* (for AP and CP, respectively) were also supported by colocalization analysis. Overall, these results expand the known genetic architecture of pancreatitis and identify tractable therapeutic targets.

## Introduction

Acute pancreatitis (AP) is a common acute inflammatory injury of the exocrine pancreas with a broad spectrum of clinical manifestations ranging from abdominal pain to multi-system organ failure and death. Overall, AP accounts for over 300,000 emergency department visits and is associated with nearly $3 billion in healthcare costs per year in the United States^1,2^. Chronic pancreatitis (CP), by contrast, is a fibroinflammatory disease producing irreversible pancreatic fibrosis, and can result in both pancreatic exocrine and endocrine insufficiency, and is a risk factor for pancreatic cancer^3^. Together, AP and CP represent a disease continuum: approximately 10% of individuals with a first episode of AP and 36% of those with recurrent episodes eventually progress to CP^4^. Both diseases share notable risk factors including alcohol use, smoking, hypertriglyceridemia, and various genetic factors. Important AP-specific causes are gallstones, which are the leading cause of AP, and mechanical instrumentation such as endoscopic retrograde cholangiopancreatography (ERCP) ^1–3,5^.

Both rare and common variants in multiple genes have been strongly associated with AP and CP, including cationic trypsinogen (*PRSS1*), cystic fibrosis transmembrane conductance regulator (*CFTR*), serine protease inhibitor Kazal type 1 (*SPINK1*), and chymotrypsin C (*CTRC*)^2,3,5–9^. Although the genetic basis of pancreatitis has been investigated extensively, genome-wide association studies (GWAS), which identify associations between common variants and disease, have been partly constrained by relatively modest numbers of cases. To date, the largest GWAS for pancreatitis was an AP meta-analysis restricted to the European (EUR) population; it included nearly 11,000 individuals with pancreatitis and identified five genome-wide significant loci^10^. For CP, the largest study to date had a discovery cohort of nearly 2,000 individuals from EUR and also identified five genome-wide significant loci^11^. However, the extent to which AP and CP share a common genetic basis, versus arising from distinct susceptibility loci, remains poorly defined.

Here, we performed a multi-population GWAS meta-analysis of both AP and CP in over 23,000 individuals with AP and over 9,000 individuals with CP across 5 biobanks and multiple populations (5 for AP and 3 for CP), to identify susceptibility loci and genes associated with pancreatitis. First, we utilized a variety of approaches to nominate putative risk genes and variants for pancreatitis. Then, we analyzed the shared and distinct genomic architecture of AP and CP at both the genome-wide and locus-specific levels. Additionally, we identified potentially causal plasma proteins and druggable proteins associated with pancreatitis. Finally, building on our gene prioritization and plasma proteome analysis, we used data from the electronic health records (EHR) of a large academic health system to test the hypothesis that there is an association between blood type and pancreatitis. Overall, we performed a large-scale genomic analysis of pancreatitis, identifying novel susceptibility loci and risk genes, and showing that blood type is a modest but significant risk factor.

## Methods

### Genome-wide association meta-analysis

We obtained GWAS summary statistics for AP and CP from UK Biobank (UKB) ^12^, BioBank Japan (BBJ) ^13^, FinnGen^14^, All of Us (AoU) ^15^, and the VA Million Veteran Program (MVP) ^16^. Acute and chronic pancreatitis were defined in a biobank-specific manner either based on Phecodes or ICD categorizations. The UKB summary statistics included individuals from European (EUR) and Central/South Asian (CSA) populations, the BBJ data included individuals from East Asian (EAS) populations, and the FinnGen data comprised individuals from EUR populations. AoU and MVP both included individuals from African (AFR), Admixed American (AMR), and EUR populations. All summary statistics underwent standard processing: 1) GWAS summary statistics from UKB and BBJ were lifted over from GRCh37 to GRCh38 coordinates using the R package rtracklayer; 2) GWASinspector was used for allele harmonization and quality control ^17^; 3) Allele count was filtered such that there were at least 50 minor alleles at each variant. Finally, we performed fixed-effects, inverse-variance-weighted GWAS meta-analyses within and across populations using METAL^18^, for both AP and CP. Independent loci were defined using the get_loci function from the R package gwasRtools, which iteratively selects the most significant variant (*P*<5×10⁻^8^) and groups all variants within ±500 kb into the same locus until all variants are assigned^19^. The nearest genes were assigned to lead variants using the get_nearest_gene function from gwasRtools^19^.

### Approximate Bayes factor fine mapping and gene prioritization

To prioritize possible causal variants at significant loci, we performed Bayesian fine mapping of the multi-population meta-analysis summary statistics, analyzing AP and CP separately. Fine-mapping was implemented using the finemap.abf function in the coloc R package, which utilizes the approximate Bayes factor (ABF) method of Wakefield et al. ^20,21^. The loci were defined as above (±500 kb region flanking each lead variant). Under the assumption of a single causal variant per locus, each variant’s posterior inclusion probability (PIP) was obtained by normalizing the ABFs across all variants within the locus, so that the PIPs, conditional on a causal variant being present in the region, summed to one. Variants were ranked by PIP and added to the credible set in decreasing order until the cumulative posterior probability reached 0.95, yielding the 95% credible set for each locus.

For gene prioritization from ABF fine-mapping, we used the Open Targets database^22^ to identify the most severe predicted consequence of each variant in the 95% credible sets. A gene was prioritized if a credible-set variant was annotated for that gene as a splice acceptor, splice donor, frameshift, missense, in-frame insertion or deletion, transcription factor binding site, or regulatory region variant. Loci whose 95% credible set contained more than 1,000 variants were excluded from this analysis, as credible-set membership provides little localization information at such loci.

### Gene prioritization using MAGMA

Gene-level association statistics were computed using MAGMA v1.10 ^23^. SNPs were assigned to protein-coding genes on the basis of NCBI build 37.3 gene locations, with variants matched between the summary statistics and the reference panel by rsID. Gene-level p-values were derived using MAGMA’s SNP-wise mean model, which aggregates the association signal of all SNPs within a gene while accounting for the local linkage disequilibrium (LD) structure. LD was estimated from the EUR subset of the 1000 Genomes Phase 3 reference panel. Genome-wide significance was defined using a Bonferroni correction for the number of genes tested (P corrected < 0.05).

### Colocalization of pancreas eQTLs and meta-analyzed pancreatitis GWAS signal

We tested for shared genetic signals between pancreatitis risk and pancreatic gene expression using coloc^21^. The analysis was performed using our meta-analysis summary statistics and pancreatic cis-eQTL summary statistics from GTEx v10^24^. Independent risk loci were defined by genome-wide-significant lead SNPs as above. At each locus we considered as candidate genes any eGene whose cis window (transcription start site ± 1 Mb) overlapped the GWAS region (lead SNP ± 500 kb). For each candidate gene, both the GWAS and eQTL data were restricted to a common analysis window centered on the GWAS lead SNP (± 250 kb), so that posterior probabilities were computed over an identical genomic span across all genes at a locus; genes for which this window extended beyond the available cis-eQTL data were excluded to avoid asymmetric regions. Within each window, GWAS effect sizes and eQTL slopes were harmonized. Colocalization was assessed with the R function coloc.abf under the single-causal-variant assumption, modeling the eQTL as a quantitative trait and the GWAS as a case/control trait, using the default prior for a shared causal variant (p12=1 × 10⁻⁵). This method calculates the posterior probabilities of five hypotheses: H0 (no causal variants for either trait), H1 (causal variant for the first trait but not the second), H2 (causal variant for the second trait but not the first), H3 (each trait has a distinct causal variant), and H4 (the two traits share a causal variant). When assessing colocalization between two traits, the hypothesis of interest is H4—with higher posterior probabilities providing stronger evidence of a shared causal variant. Default prior probabilities were used: *P*_1_=1e-4, *P*_2_=1e-4, *P*_12_=1e-5, where *P*_1_ and *P*_2_ are the prior probabilities that a variant is associated with trait 1 (pancreatic expression) and trait 2 (AP or CP), respectively. *P*_12_ is the prior probability that the variant is associated with both traits. The default prior values for *P*_1_ and *P*_2_ are generally assumed to be widely applicable^25^.

### Cross-trait genomic correlation analysis

SNP-based heritability and cross-trait genomic correlations between AP, CP, and multiple clinically related traits were estimated using linkage disequilibrium score regression (LDSC) in the R package GenomicSEM^26,27^. We restricted our analysis to EUR summary statistics to match the population of the UK Biobank–based European LD panel.

### Local genomic correlation analysis

To identify genomic regions where the genetic relationship between acute and chronic pancreatitis diverges from the genome-wide average, we performed local genomic correlation analysis using LAVA (Local Analysis of [co]Variant Association)^28^. Analyses were restricted to EUR summary statistics to match the LD reference panel. We used the LAVA-provided partition of the genome into 2,495 approximately LD-independent blocks, and a UK Biobank–based European LD reference panel. Because the two GWAS shared samples, we estimated the sampling correlation between the traits from the cross-trait LDSC intercept and provided this to LAVA to correct for sample overlap. At each locus we first performed the univariate test of local heritability for both traits, and estimated the bivariate local genomic correlation only at loci where both traits exhibited significant local heritability (Bonferroni-corrected univariate *P*<0.05/2,495). Bivariate test p-values were corrected for the number of tests performed using the Benjamini-Hochberg false discovery rate, and local genomic correlations were considered to diverge from the genome-wide estimate where their 95% confidence interval excluded the global genomic correlation (rg=0.89) estimated by LDSC on the same summary statistics as above. Analyses were conducted in R using the LAVA package (v0.1.5).

### Proteome-wide MR

We performed causal inference using two-sample MR, an approach that depends on three core assumptions: relevance (the genetic instruments are associated with the exposure), exclusion restriction (the instruments affect the outcome only through the exposure), and independence (the instruments are not associated with confounders of the exposure and outcome)^29,30^. In practice, the first assumption can be met by using variants that are significantly associated with the exposure, as determined by a GWAS. The second and third assumptions are more difficult to satisfy. However, if the exposures are gene expression levels or protein levels, one can use *cis* genetic variants—which are variants found within a fixed distance (typically 500 kb or 1 Mb) of the gene encoding the transcript or protein in question—to minimize pleiotropy and maximize biological plausibility. Additionally, directional pleiotropy can be tested using MR-Egger regression^31^. A significant intercept term could indicate directional pleiotropy and a violation of the second assumption.

For exposure data, we used publicly available pQTLs identified in two large-scale plasma-proteomics studies (deCODE and UKB-PPP) to generate genetic instruments^32,33^. The summary statistics for the deCODE dataset are available at https://www.decode.com/summarydata/, and for the UKB-PPP dataset at http://ukb-ppp.gwas.eu. For outcome data, we used our multi-population meta-analyzed GWAS summary statistics for AP and CP. Variants were chosen as instrumental variables if their nominal *P*-values achieved genome-wide significance (*P*<5e-8). Only variants present in the exposure and outcome summary statistics were considered. The instrumental variables had effect sizes for plasma pQTL levels quantified in units of standard deviations, whereas the effect sizes for pancreatitis were quantified in units of log-odds change in outcome. To reduce the risk of pleiotropic effects, we only considered *cis* variants located within 500 kb of their respective genes. To select independent variants we performed linkage disequilibrium (LD) clumping using the ld_clump function from the R package ieugwasr (version 1.0.0)^34^. To allow more instruments per protein we relaxed the maximum LD R-squared threshold to 0.1. Otherwise, default argument values were used. To ensure consistent effect direction between studies, variant harmonization was performed using reference/alternate alleles included in the summary statistics and the harmonise_data function from the R package TwoSampleMR (version 0.5.9)^35^.

We performed inverse-variance weighted MR using the mr_ivw function from the R package MendelianRandomization (version 0.10.0)^36^. In cases where there is only a single genetic variant per exposure, the mr_ivw function returns a Wald ratio. Due to our relaxed LD threshold for variants, we performed MR with the ‘correl’ argument set to ‘TRUE’ to allow for correlated instrumental variables. This adjusts the standard error of the instrumental variables to account for any residual correlation (LD) between them. The mr_ivw function returns effect size estimates for the effect of an exposure (plasma protein levels) on an outcome. Effect sizes significantly above 0 or odds ratios (ORs) significantly above 1 indicate that increased plasma levels of a protein are associated with increased genetic liability for pancreatitis. The Benjamini-Hochberg procedure was used to calculate a false discovery rate (FDR). Results were deemed significant at a threshold of FDR <0.05 in each dataset.

We then performed Bayesian colocalization between plasma protein levels and pancreatitis using the coloc.abf function from the R package coloc, as described above. We utilized all variants where summary statistics were available within 500 kb of genes encoding the proteins of interest. A posterior probability of a shared causal variant (H_4_) >0.7 was used as strong evidence of colocalization.

### Association between blood type and pancreatitis in EHR data

We used electronic health records from Penn Medicine, a large multi-hospital academic health system serving the greater Philadelphia area, to ascertain individuals with and without pancreatitis, as determined by ICD-10 code K85 for acute pancreatitis and K86.0 and K86.1 for chronic pancreatitis. We then retained individuals with blood type information. Our analysis used data from 654,946 total individuals with a known blood type, including 10,089 individuals with documented episodes of AP and 3,820 with documented CP. Association between blood type and pancreatitis was tested with logistic regression using the glm function (with “family” set to “binomial”) from the stats R package. We used the following formula: outcome ∼ blood type + sex + current age + self-reported race, where outcome was either AP or CP. Blood type had 4 possible levels: A, B, AB, and O which was considered the baseline; the Rh factor was ignored. The logistic regressions for AP and CP were performed separately.

### Drug-target Mendelian randomization

Target genes were obtained from ChEMBL^37^, with maximal clinical phase set to 1 or greater, and restricted to single-protein and protein-complex targets. Genetically predicted expression of each target was instrumented with cis-eQTLs from several sources. Whole-blood instruments were obtained from eQTLGen^38^, while instruments from pancreatitis relevant tissues (pancreas, liver, adipose (subcutaneous), adipose (omentum), terminal ileum, and spleen) were obtained from GTEx v10. Approximately independent instruments were obtained from eQTLGen using LD clumping, as described for the plasma proteomics MR. For GTEx tissues, independent instruments were obtained from published SuSiE fine-mapping results, taking the highest-PIP variant of each credible set as a conditionally independent signal. Because eQTLGen effect sizes are in standard deviation units, and GTEx fine-mapping allelic fold-change (aFC) is in log_2_ fold-change units, estimates were not compared across tissues, and were instead interpreted for direction and significance rather than magnitude. Using our multi-population meta-analyzed GWAS summary statistics for our outcome data, we performed two-sample MR as described above in the plasma proteomics section.

## Results

### Multi-population meta-analysis identifies 16 loci associated with acute pancreatitis and 14 loci associated with chronic pancreatitis

We performed a GWAS meta-analysis for acute and chronic pancreatitis using an inverse variance fixed-effects model and data from five large-scale biobanks: UK Biobank^12^, BioBank Japan^13^, FinnGen^14^, All of Us^15^, and the VA Million Veteran Program^16^. In total the meta-analyses included 23,292 individuals with and 1,748,420 individuals without acute pancreatitis (AP) and 9,866 individuals with and 1,627,720 individuals without chronic pancreatitis (CP) (**Table S1**). The AP meta-analysis spanned 5 populations (EUR, EAS, AFR, AMR, CSA), while the CP meta-analysis spanned 3 (EUR, EAS, AFR). Overall, the analysis identified 16 distinct loci associated with AP and 14 loci associated with CP at a genome-wide significance level of *P*<5×10⁻^8^ (**Figure 1, Figure S1, Table S2**). The effect sizes for each meta-analyzed lead variant within each population are shown in **Figures S2-S3**.

**Figure 1.**
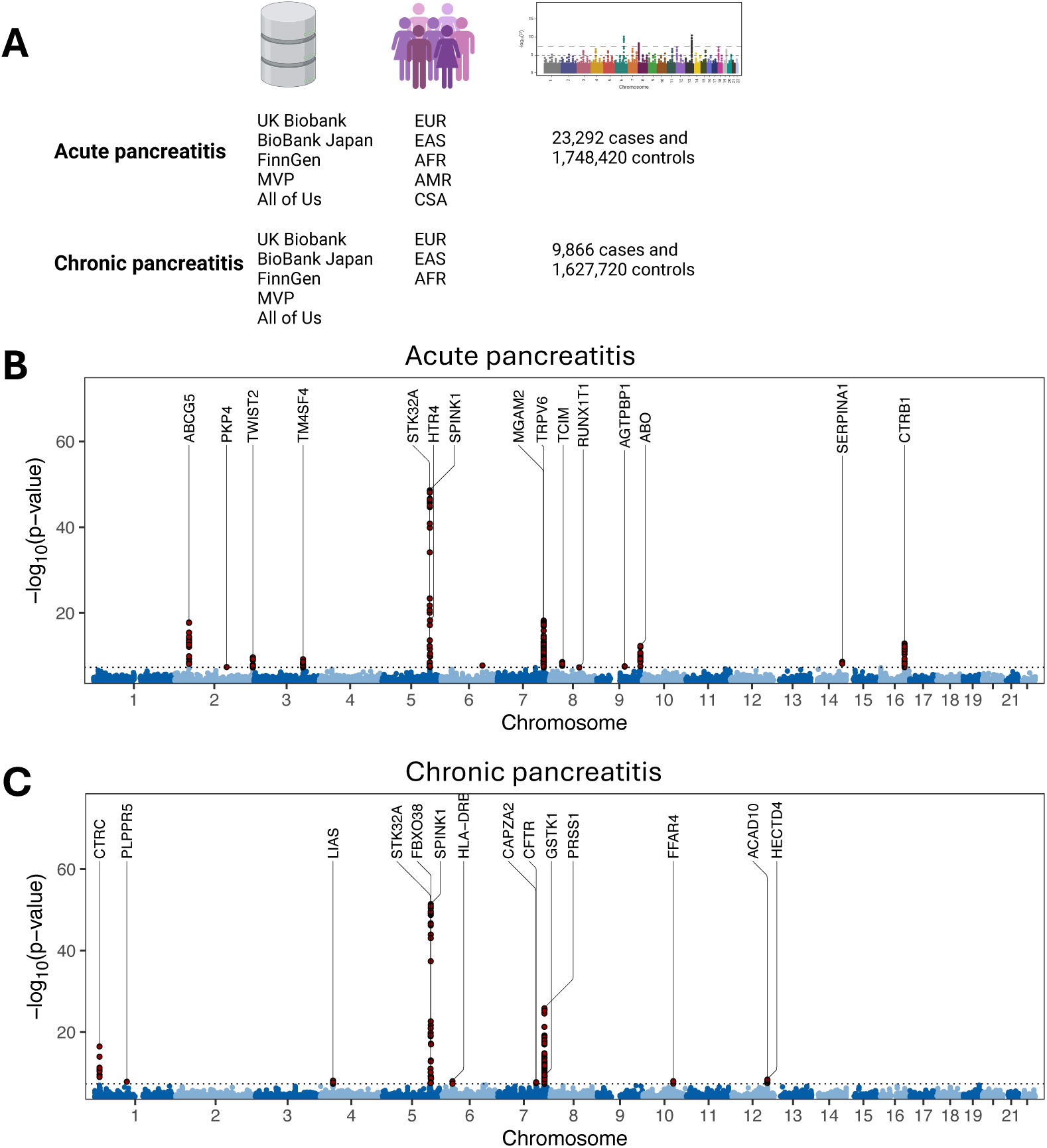
Multi-population GWAS meta-analysis of acute and chronic pancreatitis. (A) Overview of the biobanks and populations included in the meta-analysis, with total case and control counts. Manhattan plots of the inverse-variance-weighted fixed-effects meta-analysis for (B) acute and (C) chronic pancreatitis. Red dots indicate genome-wide significant variants (*P*<5×10^-8^). Nearest genes to each significant locus are labeled.

### Fine mapping and functional annotation prioritize genes associated with pancreatitis

To refine the set of variants associated with pancreatitis across all significant loci, we fine-mapped our multi-population meta-analysis results using approximate Bayes factor (ABF) analysis^39^. The 95% credible sets ranged from 1 to 17,136 variants for AP (median 12 per locus) and from 1 to 14,924 variants for CP (median 11 per locus) (**Table S3**). Resolution varied widely across loci: while most signals fine-mapped to compact sets, a minority resolved poorly, consistent with the presence of multiple causal signals or population-divergent LD at those loci (3/16 AP and 3/14 CP loci contained more than 1,000 variants). Because credible-set membership is uninformative for prioritization at such loci, these were carried forward through the other prioritization approaches but excluded from the credible-set–overlap analysis.

To prioritize genes from our multi-population meta-analyses, we first identified genes containing candidate functional variants that overlapped the fine-mapping credible sets (**Figure 2A**); this approach prioritized 4 genes for AP and 5 for CP. We applied three additional prioritization approaches (**Figure 2A**): assignment of the nearest gene to the index variant at each locus, gene-level MAGMA analysis^23^, and colocalization between pancreas expression quantitative trait loci (eQTLs) and the meta-analyzed GWAS signals^21^. MAGMA identified 8 genes for AP and 6 for CP at a Bonferroni-adjusted *P*<0.05 (**Table S4**). Finally, the pancreas eQTL–GWAS colocalization analysis identified 7 genes for AP and 4 for CP whose pancreatic eQTL signals showed a colocalization posterior probability (PP.H4) greater than 0.7 with one of the GWAS peaks (**Table S5**). Overall, 7 genes were prioritized for AP through 2 or more approaches: *ABCG5*, *ABO*, *CTRB1*, *SPINK1*, *TCIM*, *TM4SF4*, and *TWIST2*. Similarly, 5 genes were prioritized for CP through 2 or more methods: *CTRC*, *FFAR4*, *HLA-DRB1*, *PRSS1*, and *SPINK1*. Several of the prioritized genes across both phenotypes, namely *PRSS1*, *SPINK1*, *CTRC*, and *CTRB1*, encode components of the trypsin activation/inhibition pathway and are known to be involved in the genetics of pancreatitis^5,9^. Additionally, *TWIST2* and *ABCG5* were implicated in a recent GWAS meta-analysis for acute pancreatitis^10^ and *HLA-DRB1* has been previously implicated in autoimmune pancreatitis, a subtype of chronic pancreatitis^40,41^. *ABO* has been associated with chronic pancreatitis but not previously with acute pancreatitis^42^, though it is a highly pleiotropic locus. To our knowledge *TCIM*, *TM4SF4*, and *FFAR4* are novel associations for pancreatitis. The locus plots for *TM4SF4* and *FFAR4* for pancreas eQTL data and our GWAS summary statistics are shown in **Figure 2B**. For *TM4SF4* the most significant GWAS variant found in both eQTL and AP GWAS data was rs4482615 (GRCh38, chr3:149480894 T/C) with the T allele being associated with increased pancreatic expression of *TM4SF4* (*β*=0.51) and increased risk for AP (OR=1.06). For *FFAR4* the most significant GWAS variant at the locus was rs9630085 (GRCh38: chr10:93573306 A/G) with the A allele associated with increased pancreatic expression of *FFAR4* (*β*=0.80) while also being associated with decreased CP risk (OR=0.91).

**Figure 2.**
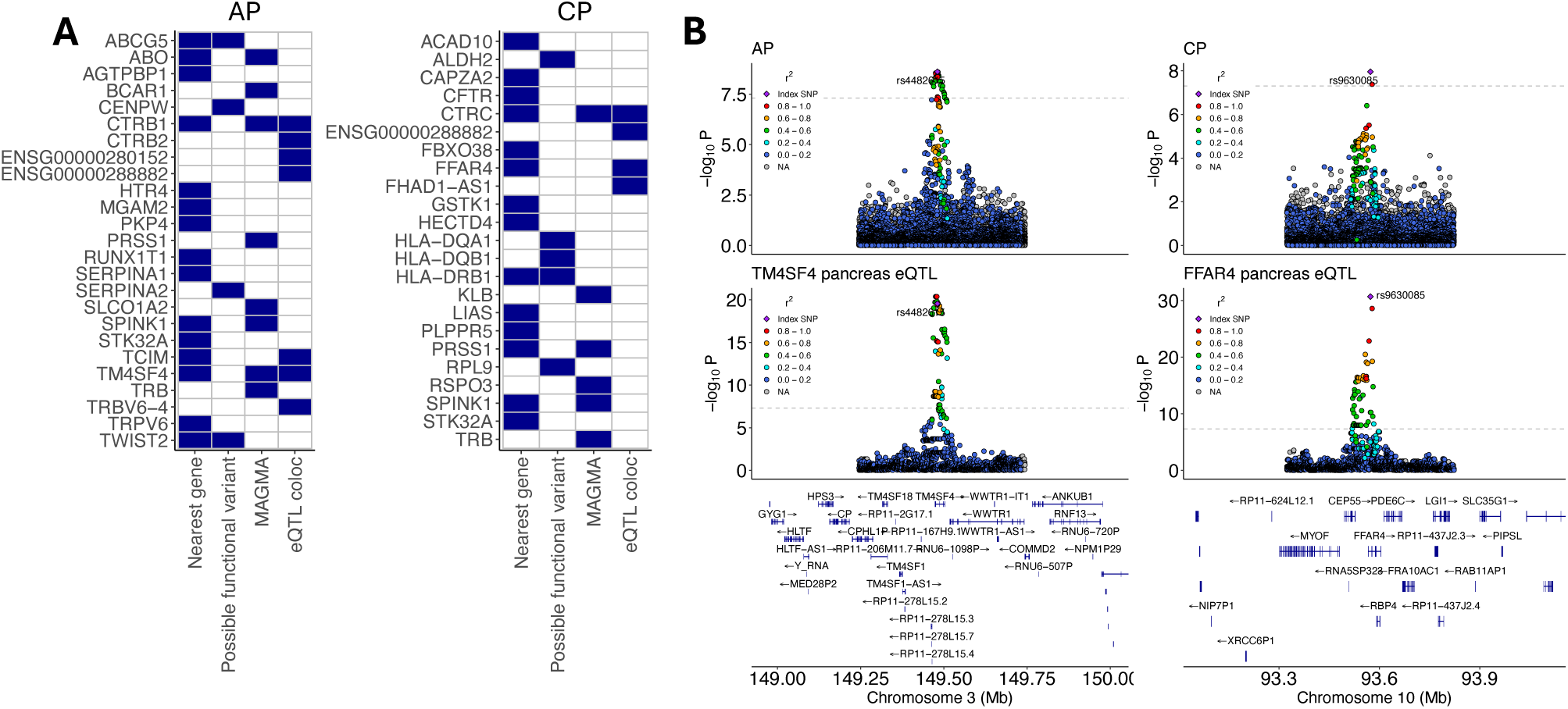
Gene prioritization, fine-mapping, and colocalization. (A) Summary of gene prioritization. The nearest gene was defined as the closest gene to the lead variant at each locus. Genes with a possible functional variant were identified from 95% credible sets from ABF fine-mapping, together with whether the variant was predicted to be functional according to Open Targets. Genes were considered significant by MAGMA if the adjusted *P*-value was < 0.05 in the gene-level MAGMA analysis. Finally, colocalization analysis between pancreas eQTLs and the meta-analyzed GWAS signal was performed to identify genes with a posterior probability of colocalization (PPH4) greater than 0.7. (B) Locus plots for two genes whose eQTLs colocalized with the meta-analyzed pancreatitis GWAS signal. In pancreatic tissue, the *TM4SF4* eQTL colocalized with the AP GWAS signal (PPH4 = 0.98) and the *FFAR4* eQTL colocalized with the CP GWAS signal (PPH4 > 0.99).

### Shared genetic architecture between pancreatitis and related traits

We next examined the shared genetic architecture between AP and CP, and between pancreatitis and related traits. Using linkage disequilibrium score regression (LDSC), we estimated liability-scale SNP heritability at 0.047 (SE=0.0062) for AP and 0.045 (SE=0.0065) for CP, assuming population prevalences of 1.5% and 0.15%, respectively.

We then estimated genome-wide genomic correlations r_g_ among AP, CP, and several clinically related traits (**Figure 3A**). AP and CP were strongly correlated (r_g_=0.89, SE=0.11). Both traits also correlated with their established precipitants, though to differing degrees: AP showed correlations of 0.66 (SE=0.07) with gallstones, 0.42 (SE=0.05) with triglycerides, and 0.17 (SE=0.04) with alcohol use, whereas the corresponding estimates for CP were 0.41 (SE=0.07), 0.30 (SE=0.05), and 0.32 (SE=0.05). Additionally, AP showed a moderate correlation with type 2 diabetes (rg=0.34, SE=0.04), whereas CP showed a weaker correlation (rg=0.19, SE=0.05). Neither AP nor CP was substantially correlated with type 1 diabetes (AP: rg=0.08, SE=0.06; CP: rg=0.13, SE=0.06).

**Figure 3.**
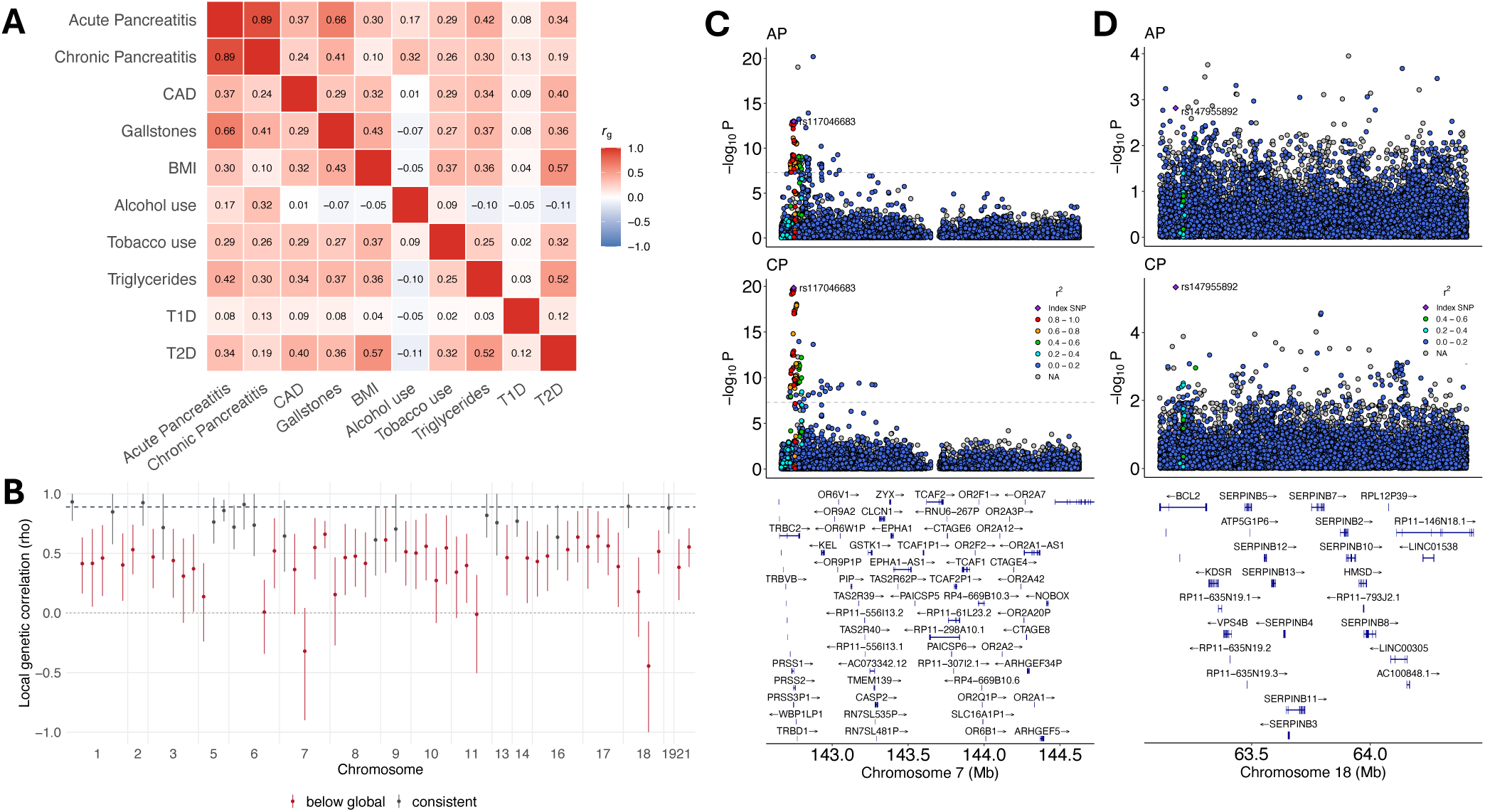
Genomic correlations between pancreatitis and clinically related traits. (A) Heatmap of genomic correlations between AP, CP, and other clinically related traits. (B) Results of local genomic correlation analysis from LAVA. Fifty-one independent regions with significant heritability in both AP and CP were tested for local genomic correlation between the two traits; regions colored in red had 95% confidence intervals that did not intersect the global genomic correlation of 0.89 (upper dashed line). (C) Region on chromosome 7 containing *PRSS1* and *PRSS2*, which has a significantly lower local genomic correlation (ρ = 0.66, 95% CI 0.54–0.77) than the global correlation. (D) Region on chromosome 18, containing *BCL2*, with a significant negative correlation between AP and CP (ρ = −0.44, 95% CI −1.00 to −0.07).

Given that AP and CP share largely the same genetic architecture when averaged across the genome, we next examined local genomic correlations between the two traits to identify regions of divergence. Using LAVA^28^ we identified 51 regions with significant non-zero local genomic correlations between AP and CP at FDR<0.05 (**Table S6**). Separately, we identified 44 regions where the 95% confidence interval of the local genomic correlation excluded the global genomic correlation (0.89; **Figure 3B**); in all 44, the local correlation was lower than the global estimate. Two of these 44 regions contained a genome-wide significant peak (*P*<5×10⁻⁸) for at least one of the traits, with one of them notably containing the *PRSS1-PRSS2* locus (rho=0.66, 95% CI 0.54-0.77) (**Figure 3C**). Among the divergent regions, one on chromosome 18 showed a significantly negative local genomic correlation (rho=−0.44, 95% CI −1.00 to −0.07) (**Figure 3D**). Although that region did not contain any genome-wide significant loci, it showed significant heritability in both AP and CP (h^2^ on observed scale=0.0035, *P*=1.41×10⁻^5^; h^2^=0.0021, *P*=3.12×10⁻^8^, for CP and AP respectively) and contains the well-known anti-apoptotic gene *BCL2* and multiple clade B serpin genes, which are a group of intracellular protease inhibitors that protect cells from proteolysis and regulate inflammation and apoptosis^43^.

### Proteome-wide Mendelian randomization reveals associations between plasma proteins and pancreatitis

To identify possible causal associations between plasma protein levels and pancreatitis, we used two-sample Mendelian randomization (MR) to estimate the effects of genetically determined plasma protein levels on the genetic liability for acute pancreatitis (AP) and chronic pancreatitis (CP). Using two-sample MR with inverse-variance weighting, we tested a total of 2,690 and 2,755 unique plasma proteins for associations with AP and CP, respectively. These comprised 1,575 (AP) and 1,680 (CP) proteins from the deCODE plasma proteomics dataset^32^ and 1,924 (AP) and 1,925 (CP) proteins from the UK Biobank (UKB) dataset^33^. The number of genetic variants used as instruments per protein ranged from 1 to 172.

We identified 15 and 6 plasma proteins associated with AP and CP, respectively, at FDR < 0.05 (**Figure 4, Table S7**). Notable positive associations included *ABO*, for which increased genetically predicted levels were associated with higher risk of both AP and CP (OR=1.04, *P*=1.15×10⁻²² for AP; OR=1.09, *P*=3.64×10⁻⁴¹ for CP), as well as *GCKR* (OR=1.73, *P*=1.89×10⁻⁶ for AP), MSRA (OR=2.07, *P*=5.35×10⁻⁶ for AP), and *PRSS2* (OR=1.99, *P*=2.66×10⁻⁵ for CP). Conversely, increased genetically predicted levels of several proteins were associated with reduced pancreatitis risk, including *CTRC*—a protein known to be protective against pancreatitis^5,8^ —for both AP and CP (OR=0.88, *P*=3.43×10⁻⁵ for AP; OR=0.71, *P*=7.28×10⁻¹³ for CP).

**Figure 4.**
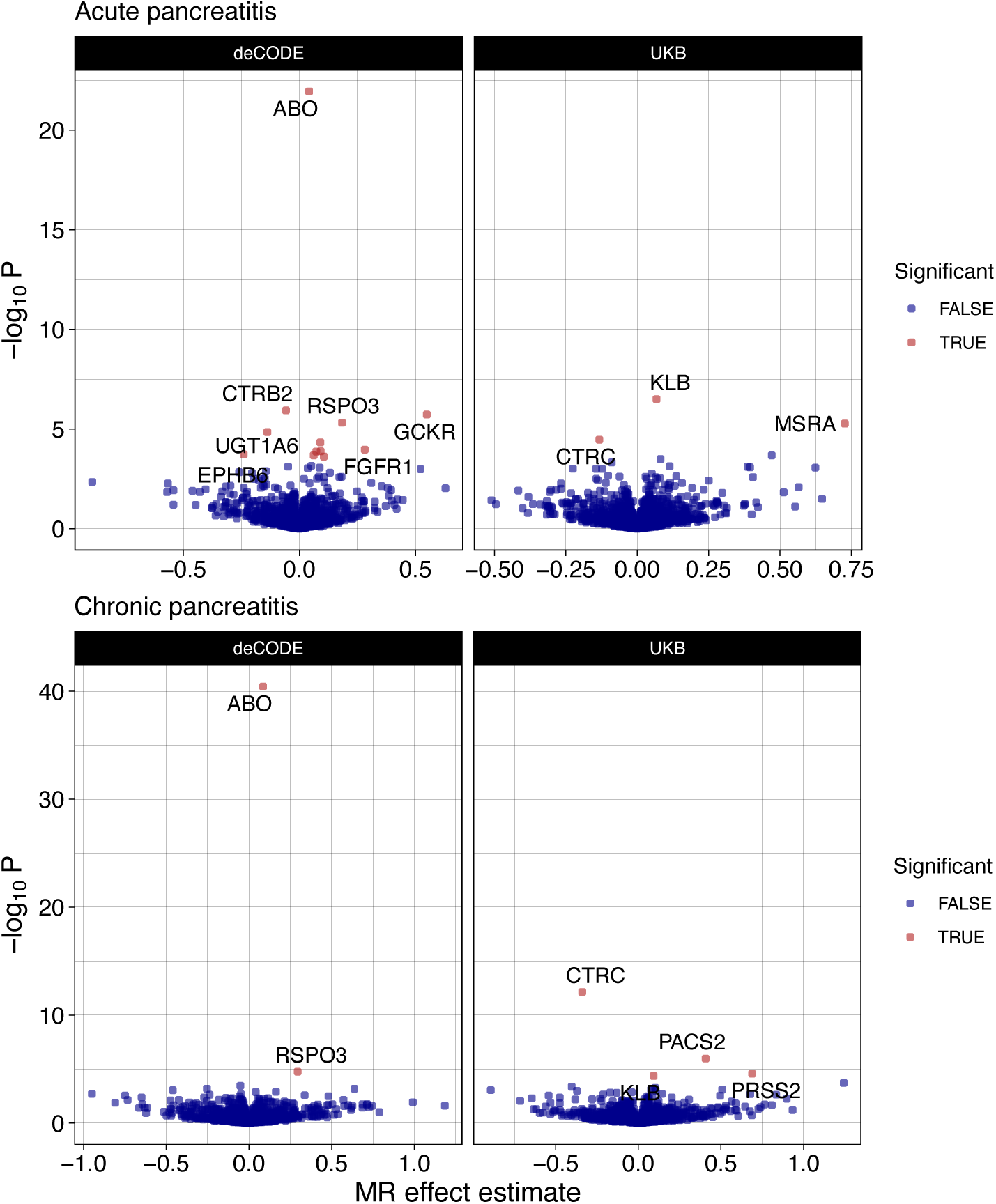
Proteome-wide MR. Proteome-wide Mendelian randomization (MR) revealed significant associations between genetically predicted plasma protein levels and pancreatitis. Overall, 15 and 6 plasma proteins were significantly associated with AP and CP, respectively. The effect estimate is the natural logarithm of the odds ratio (OR). Significance was determined at a false discovery rate (FDR) < 0.05 in each dataset.

To provide additional support for the associations between genetically determined plasma protein levels and genetic liability to pancreatitis, we tested for evidence of colocalization between protein quantitative trait loci (pQTLs) and their cognate pancreatitis risk loci. This approach helps distinguish putative causal associations from those confounded by linkage disequilibrium ^44^. Colocalization was performed using all cis variants within 500 kb of their respective genes. We identified 5 proteins for each of AP and CP with evidence of colocalization between the pQTL signal and the GWAS meta-analysis signal (posterior probability of H4 > 0.7; **Figure S4**, **Table S8**).

Notably, ABO was prioritized for pancreatitis as a nearest gene (for AP), through MAGMA (for AP), and through MR in the deCODE proteomics dataset (for both AP and CP). However, ABO is a known pleiotropic locus making MR interpretation challenging^45^. To better understand whether directional horizontal pleiotropy affected our MR results we performed MR-Egger. We found that for both AP and CP the small but significant MR effect was preserved and the intercept, which is a test for directional horizontal pleiotropy, was not statistically significant (OR=1.05, *P*=1.64×10⁻^14^, intercept *P*=0.33 for AP; OR=1.10, *P*<0.001, intercept *P*=0.065 for CP), suggesting that directional pleiotropy did not play a large role in the original MR results. In the deCODE plasma proteomics dataset, the aptamer for *ABO/BGAT* binds to amino acids 38-354^32^, while the most common O allele is truncated at 117 amino acids^46,47^. Therefore, it is plausible that elevated plasma levels of *ABO/BGAT* as measured by the aptamer reflect, to a certain extent, non-O-type blood.

### Electronic health records show an association between non-O blood type and pancreatitis

To further explore the relationship between pancreatitis and blood type, we examined the association between blood type and pancreatitis in electronic health records from Penn Medicine, a large multi-hospital academic health system serving the greater Philadelphia area. We used data from 654,946 total individuals with a known blood type, including 10,089 individuals with documented episodes of AP and 3,820 with documented CP ascertained from ICD-10 codes (**Table S9**). To determine whether pancreatitis was associated with blood type, we performed separate logistic regressions for AP and CP while controlling for current age, sex, and self-reported race. We found that type A blood was significantly associated with chronic pancreatitis (OR 1.13 relative to type O blood, *P*=1.16×10^-3^) and marginally associated with acute pancreatitis (OR=1.05, *P*=0.043) (**Figure 5, Table S10**). Type B blood was significantly associated with both chronic and acute pancreatitis (OR=1.14, *P*=2.75×10^-3^ for CP; OR=1.20, *P*=7.58×10^-11^ for AP). Similarly, type AB blood was also significantly associated with both chronic and acute pancreatitis (OR 1.20, *P*=1.65×10^-2^ for CP; OR 1.17, *P*=1.17×10^-3^ for AP). The associations of blood type with AP and CP, stratified by ICD code, are shown in **Figure S5**.

**Figure 5.**
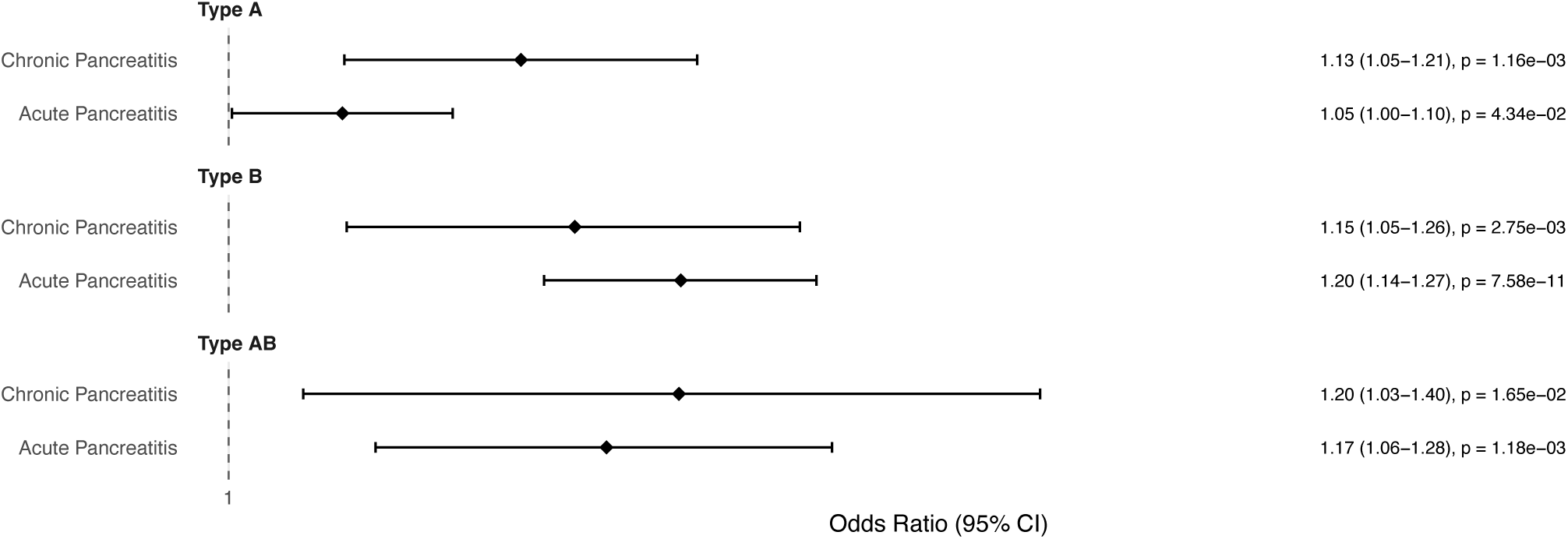
Association between blood type and pancreatitis. Blood type and diagnosis of acute and chronic pancreatitis were obtained from electronic health records from Penn Medicine. Logistic regressions were performed for AP and CP, controlling for current age, sex, and self-reported race. The reported odds ratios are relative to type O blood. Confidence intervals are 95% confidence intervals.

### Drug repurposing mendelian randomization identifies possible drug targets associated with pancreatitis

Whereas the proteome-wide analysis surveyed circulating plasma proteins, we next sought to identify druggable targets amenable to therapeutic repurposing by testing genetically determined gene expression across disease-relevant tissues. We restricted this analysis to clinical-stage drug targets and used two-sample MR to estimate the effects of genetically predicted expression of each druggable target on the genetic liability for AP and CP. Instruments comprised conditionally independent *cis*-eQTLs from whole blood (eQTLGen; 594 targets) and six pancreatitis-relevant GTEx tissues (pancreas, liver, subcutaneous and visceral adipose, terminal ileum, and spleen; 366 targets), and we tested 690 unique druggable target genes for each of AP and CP, corresponding to 1,279 (AP) and 1,281 (CP) gene–tissue associations. The number of genetic variants used as instruments per target ranged from 1 to 128.

We identified 38 and 45 druggable target genes associated with AP and CP, respectively, at FDR < 0.05 (**Figure 6, Table S11**). Notable positive associations included DRD2, for which increased genetically predicted expression in adipose tissue was associated with higher risk of AP (OR=1.06, *P*=2.72×10^-4^ in visceral adipose; OR=1.06, *P*=3.59×10^-4^ in subcutaneous adipose), as well as *CXCR2* (OR=1.10, *P*=3.01×10^-12^ in whole blood) and *AKT1* (OR=1.26, *P*=5.32×10^-6^ in pancreas) for AP, and *BCHE* (OR=1.07, *P=*6.90×10^-5^ in subcutaneous adipose) for CP. Conversely, increased genetically predicted expression of several targets was associated with reduced pancreatitis risk, including *IL18R1* for both AP and CP (OR=0.96, *P*=1.82×10^-5^ for AP; OR=0.88, *P*=7.92×10^-16^ for CP), consistent with a shared contribution of interleukin-18 receptor signaling to both subtypes.

**Figure 6.**
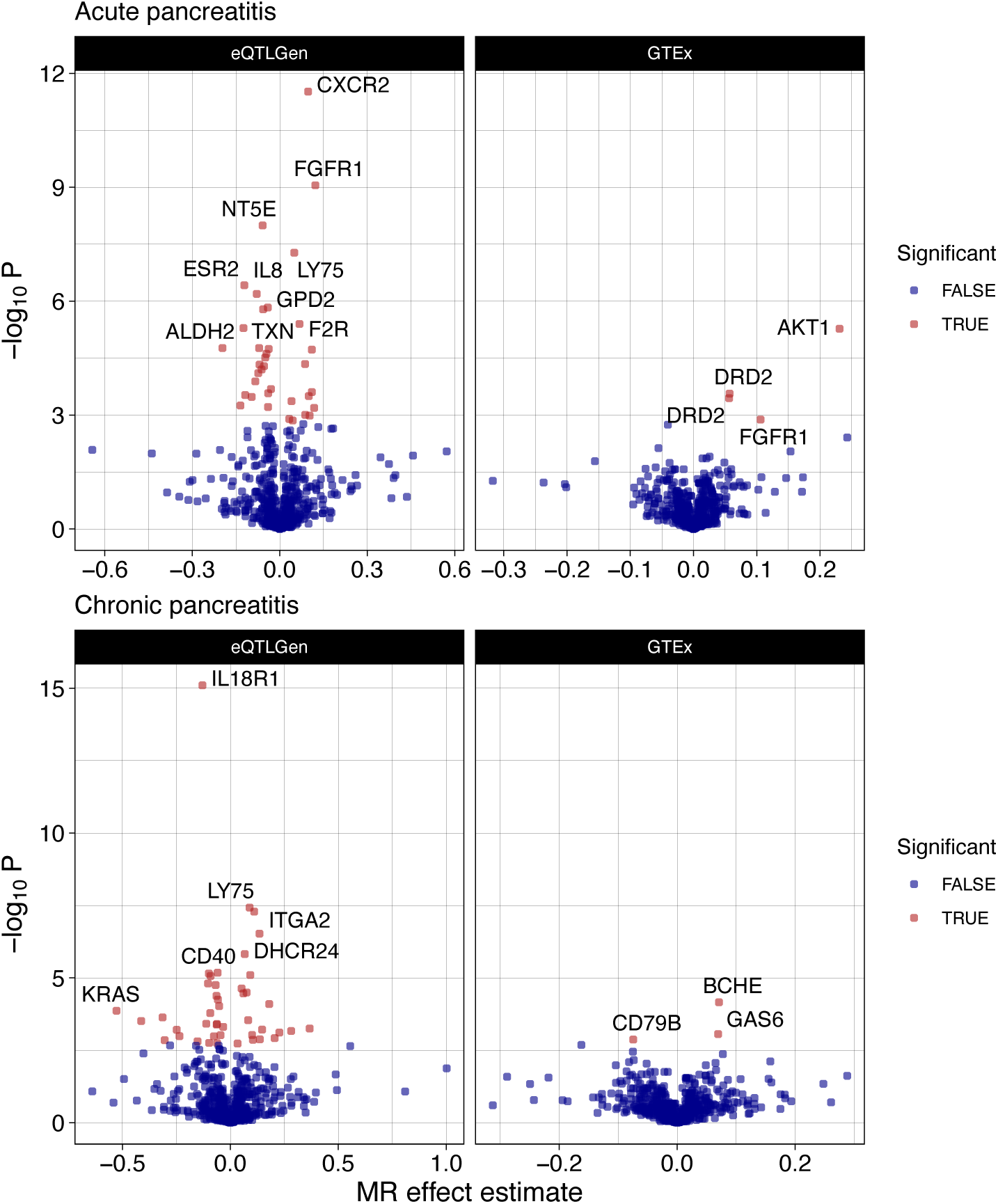
Drug-target MR. MR revealed significant associations between genetically predicted whole-blood and tissue mRNA levels of druggable genes and pancreatitis. Overall, 38 and 45 druggable genes were significantly associated with AP and CP, respectively. The effect estimate is the natural logarithm of the odds ratio (OR). Significance was determined at a false discovery rate (FDR) < 0.05.

To provide additional support for the associations between genetically determined expression and genetic liability to pancreatitis, and to distinguish putative causal targets from those confounded by linkage disequilibrium, we tested for evidence of colocalization between each *cis*-eQTL and its cognate pancreatitis risk locus, as described above. We identified one target for each of AP and CP with evidence of colocalization (posterior probability of H4 > 0.7): the dopamine D2 receptor *DRD2* in visceral adipose tissue for AP (PP.H4=0.72, with concordant support in subcutaneous adipose, PP.H4=0.69), and butyrylcholinesterase *BCHE* in subcutaneous adipose tissue for CP (PP.H4=0.89). Both targets are the subject of approved drugs, nominating them as candidates for repurposing.

## Discussion

We performed a large multi-population GWAS meta-analysis of acute and chronic pancreatitis (AP and CP), comprising 23,292 AP and 9,866 CP cases across five biobanks, and identified 16 and 14 genome-wide significant risk loci, respectively. To prioritize candidate genes at these loci, we applied four complementary approaches and identified 7 genes for AP and 5 for CP that were supported by two or more approaches, including 3 potentially novel gene-pancreatitis associations: *TM4SF4* and *TCIM* for AP and *FFAR4* for CP. We found that AP and CP shared a genomic correlation of 0.89; however, dozens of genomic regions were identified with significantly lower local correlations, including one region containing *BCL2* with a negative correlation. Proteome-wide MR analysis identified 15 and 6 plasma proteins whose genetically predicted plasma levels were significantly associated with AP and CP, respectively. Among them was ABO, which was reasoned to be a proxy for non-O blood type and confirmed as a modest risk factor for pancreatitis in the health records of over 650,000 individuals. We also performed drug-repurposing MR and identified 38 and 45 druggable target genes associated with AP and CP, respectively. These findings carry several implications. First, genetic susceptibility for pancreatitis extends beyond the well-characterized trypsin pathway. Second, AP and CP largely share a genomic architecture; however, regions of discordance between the two traits possibly harbor therapeutic consequences. Third, non-O blood type is a modest but significant risk factor for pancreatitis. Finally, several druggable proteins are now candidates for experimental testing.

Our gene prioritization results both confirm and extend the protease-driven model of pancreatitis pathogenesis. For both AP and CP, we were able to recover the well-characterized genes of the trypsin pathway: notably *SPINK1* and *CTRB1* for AP and *CTRC*, *PRSS1*, and *SPINK1* for CP^2,3,6–8,11^. Other previously identified loci for *TWIST2* and *ABCG5* were also nominated using two complementary approaches for AP^10^. *CFTR*, a gene classically associated with chronic pancreatitis^5,9^, was supported by nearest-gene assignment for CP. Concordantly, our proteome-wide MR analysis also nominated several members of the trypsin pathway including *PRSS2* and *CTRC.* Using colocalization with pancreas eQTL data, we showed increased *FFAR4* expression was associated with decreased risk of CP. Notably, *FFAR4* is a free fatty-acid receptor which senses medium-and long-chain fatty acids and has been implicated as protective against fibrosis in multiple organs^48^. *FFAR4* has been shown to protect against renal, hepatic and cardiac fibrosis via the inhibition of profibrotic *TGF-β*1 signaling and the inhibition of proinflammatory cytokines such as *IL6* and *TNFα*. Additionally, *FFAR4* has been implicated as part of an anti-inflammatory pathway in rat pancreatic cells^49,50^. Given that CP is considered to be a disease of pancreatic inflammation and fibrosis^3^, it is biologically plausible that variants that increase pancreatic *FFAR4* levels also confer some protection against CP. Interestingly, *FFAR4* has also been implicated in pancreatic cancer, for which CP is a well-known risk factor, and has been shown to inhibit the growth of cancer while simultaneously promoting metastasis^51^. *TM4SF4* is a cell-surface protein involved in endocrine pancreatic development; however, there is no clear evidence linking it to inflammation or pancreatitis^52^. Finally, *TCIM* is a positive upstream regulator of *Wnt/β-catenin* signaling and an amplifier of the pro-inflammatory *NF-κB* pathway, but has not been linked to pancreatic pathology^53,54^. Together, our novel gene-pancreatitis associations implicate lipid sensing, inflammatory signaling, and pancreatic development and provide starting points for future functional work.

We found that AP and CP shared a genomic correlation of 0.89, which highlights the strong genetic similarity between AP and CP, and reinforces a genetic basis for the AP to CP continuum. However, local genomic correlation analysis identified dozens of genomic regions with genetic correlations significantly lower than the genome-wide average of 0.89, including one region with a significant negative correlation. The region with a negative correlation between AP and CP includes the anti-apoptotic gene *BCL2*. Notably, in mouse models of AP, *BCL2* inhibitors (venetoclax and navitoclax) have been shown to shift pancreatic acinar cells from necrosis into apoptosis and reduce disease severity. However, *BCL2* inhibition also exacerbated fibrosis in mouse CP models^55^. These experimental results align with our observation of a negative local genomic correlation at the *BCL2* locus. This region also contains multiple clade B serpin genes, intracellular protease inhibitors that interact with the same cell-death programs as BCL2, so we cannot attribute the divergent signal to BCL2 alone^43^. Thus, a high global genomic correlation cannot be used to blindly carry over treatments between AP and CP, and candidate therapies should be evaluated separately in AP and CP.

Our ABO results add multiple layers of orthogonal evidence for an association that has proven inconsistent in prior studies. For both AP and CP, we found that genetically increased plasma levels of ABO were associated with increased pancreatitis risk. Furthermore, this was supported by evidence of colocalization between the GWAS signal and the ABO pQTL for CP. This is in addition to ABO being prioritized for AP using the nearest gene approach and MAGMA. Because the deCODE aptamer does not distinguish the truncated O allele, we reasoned that increased plasma *ABO* levels were essentially acting as proxies for non-O blood type. Using Penn Medicine EHR data we found that non-O blood type was significantly associated with both AP and CP, with modest effect sizes. Prior studies have shown an inconsistent relationship between blood type and pancreatitis. A German genetic association study reported that blood type B and FUT2 non-secretor status are associated with elevated serum lipase activity and increased risk of chronic pancreatitis^42^, whereas the earlier North American Pancreatitis Study 2 (NAPS2) found blood group O to be more prevalent among chronic pancreatitis patients^56^, and a recent meta-analysis concluded that ABO and FUT2 play at most a limited role in chronic pancreatitis risk in Chinese cohorts^57^. Our study integrated GWAS, proteomic, and independent epidemiological data which consistently demonstrated an association between non-O type blood and pancreatitis. The effect sizes are too small to be usefully predictive in individual patients; however, blood type is easily ascertained and may warrant inclusion in composite risk models and should be captured prospectively in pancreatitis cohorts.

Our drug-target MR analysis revealed multiple therapeutic hypotheses which are testable with drugs that already exist. We found that increased genetically predicted *DRD2* expression in adipose tissue was associated with increased genetic liability for AP, and this was supported by colocalization analysis. Experimental evidence suggests that *DRD2* signaling in pancreatic tissue is associated with decreased inflammation and is protective against AP^58^. However, in adipose tissue, experimental evidence indicates that *DRD2* signaling is pro-inflammatory and increases *IL6* and TNF-*α* expression^59^, providing a plausible mechanistic bridge between increased *DRD2* signaling and AP. Prochlorperazine is a *DRD2* antagonist which is widely used for its effects on the central nervous system^60^. However, it is unclear whether it has more substantial effects in the pancreas or in adipose tissue. Therefore, future studies should assess its relative effects on pancreatic versus adipose *DRD2*, and ultimately whether it can be used to prevent or treat AP. Among other AP targets was *CXCR2* in whole blood. Experimental evidence in mice has shown that *CXCR2* inhibition reduced the transmigration of neutrophils and protease activation in acute pancreatitis and reduced disease severity^61^. Future studies should examine whether *CXCR2* inhibitors like ladarixin^62^ can be used to treat AP. For CP we found that increased genetically predicted *BCHE* expression in subcutaneous adipose tissue was significantly associated with increased liability for CP. This result was also supported by colocalization between subcutaneous adipose eQTL data and CP GWAS signal. *BCHE* is an enzyme in adipose tissue which hydrolyzes acetylcholine and long-chain acylcholines that normally inhibit macrophage cytokine production^63^. Consistent with this, higher *BCHE* activity in humans tracks with obesity, dyslipidemia, and low-grade inflammation^64^. Therefore, it is plausible that increased adipose *BCHE* expression exacerbates pancreatic inflammation and fibrosis leading to CP. However, this is purely a hypothetical connection and future research should examine mechanistic links between adipose *BCHE* expression and CP. Both targets supported by colocalization were identified using eQTLs from adipose tissue. This suggests that adipose inflammation could be a modifiable extrinsic contributor to pancreatitis.

These results should be interpreted in light of several limitations. First, although we performed a multi-population meta-analysis, the majority of the individuals in this study are genetically similar to European reference panels. Furthermore, our genomic correlation analysis focused solely on EUR individuals. Future analyses should include a higher proportion of individuals from diverse populations. Another substantial limitation of our study is that AP and CP are defined per biobank according to ICD categorizations and Phecodes, which conflates etiologies (such as gallstones and alcohol-associated pancreatitis), severity, and recurrence (acute vs recurrent acute pancreatitis), and is prone to AP/CP misclassification. We also relied on bulk tissue eQTL data which limits our ability to identify cell-type-specific signals related to disease. MR analysis intrinsically has multiple limitations including two assumptions that are untestable: (1) that the instrumental variables affect the outcome only through the exposure; and (2) that the instrumental variables are not associated with confounders affecting both the exposure and outcome. Also, genetically predicted tissue expression and plasma protein levels are simply proxies for protein activity and do not necessarily reflect the effects of pharmacologic manipulation. Additional experimental evidence is therefore necessary to prove causality and to confirm whether the potential drug targets identified in this study are genuine. Our ABO pancreatitis analysis carries several additional caveats. First, ABO is among the most pleiotropic loci in the genome, associated with von Willebrand factor, thrombotic and cardiovascular traits, inflammatory markers, and circulating lipids^45^. Although MR-Egger showed no strong evidence of directional pleiotropy, the breadth of these associations means a pancreas-intrinsic causal effect cannot be firmly established from these data alone. Second, because dyslipidemia and other established pancreatitis precipitants are themselves genetically correlated with AP and CP in our analyses, part of the ABO signal may act through these upstream factors rather than the pancreas directly. Third, the EHR analysis is observational, limited to individuals with a clinically documented blood type, and subject to ascertainment bias. The analysis may also retain residual confounding by genetic ancestry despite adjustment for self-reported race, as ABO allele frequencies vary across populations^65^.

In summary, we have performed a multi-population pancreatitis GWAS meta-analysis. We have identified novel risk loci and genes for pancreatitis, and examined the shared and distinct genomic architectures of AP and CP. We also identified non-O type blood as a modest but significant risk factor for pancreatitis using multiple lines of evidence. Finally, we identified possible drug targets for pancreatitis. Overall, our study supplies a concrete set of leads for two diseases with no approved disease-modifying therapies: a short list of prioritized genes, an easily ascertainable risk factor, and candidate drugs for repurposing that can be tested in existing models.

## Availability of data and materials

Data is available in the manuscript and supplementary materials. Code will be provided upon reasonable request to the authors.

## Acknowledgements

We thank the participants of the UK Biobank, FinnGen, the All of Us Research Program, BioBank Japan, and the Million Veteran Program, whose contributions made this work possible. We also thank the National Institutes of Health’s All of Us Research Program for making available the data examined in this study.

