## Supplementary Figures for "Multi-population genome-wide association meta-analysis of acute and chronic pancreatitis"

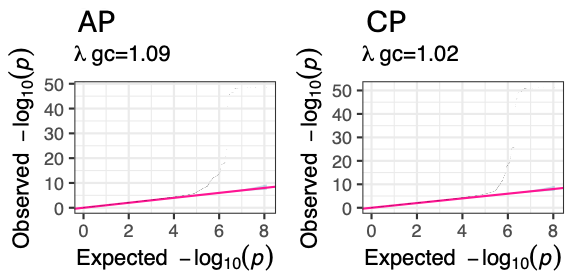

**Figure S1.** Quantile-Quantile (Q-Q) plots for both the AP and CP multi-population meta-analysis with genomic inflation for each shown.

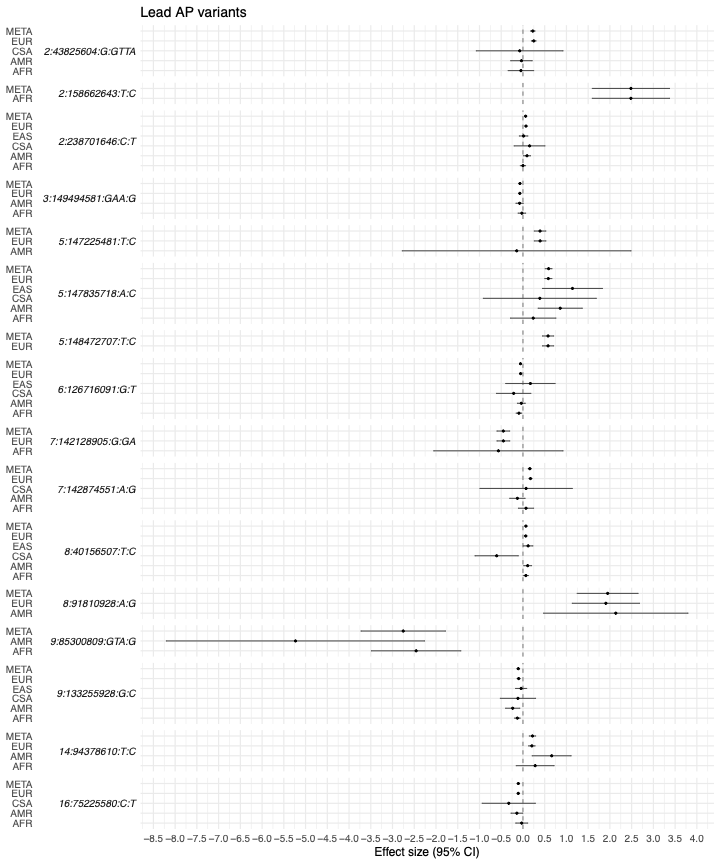

**Figure S2.** Effect sizes for the lead variants from the AP meta-analysis across populations comprising the meta-analysis. 95% confidence intervals are shown.

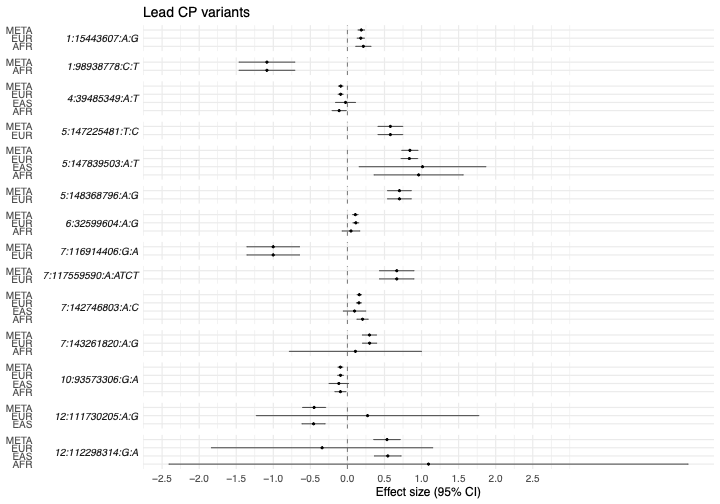

**Figure S3.** Effect sizes for the lead variants from the CP meta-analysis across populations. 95% confidence intervals are shown.

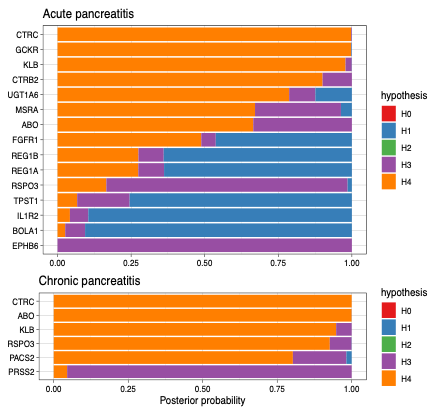

**Figure S4.** Bayesian colocalization results for all plasma proteins significantly associated with AP or CP. The hypotheses are as follows: H0, no causal variant for either trait; H1, causal variant for plasma protein levels only; H2, causal variant for pancreatitis only; H3, two distinct causal variants; and H4, one shared causal variant for plasma protein levels and pancreatitis.

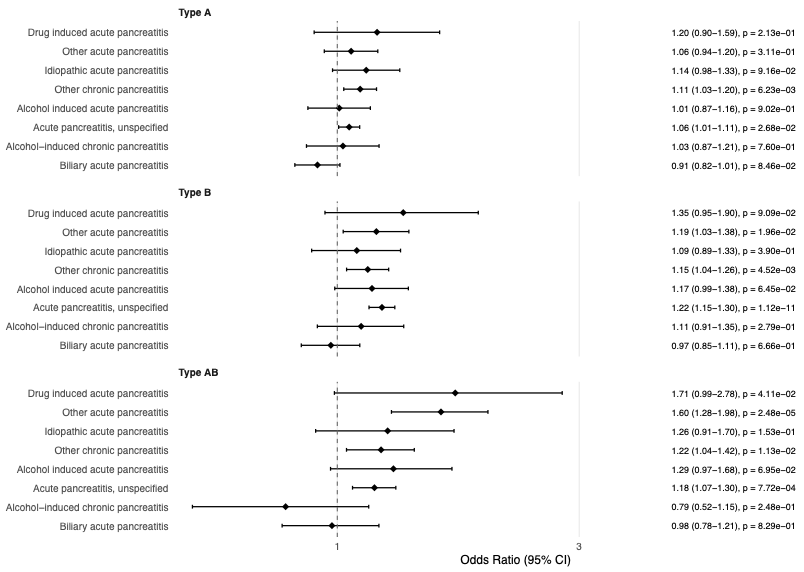

**Figure S5. Association between blood type and pancreatitis stratified by ICD code.** Blood type and diagnosis of acute and chronic pancreatitis were obtained from electronic health records in Penn Medicine. Logistic regressions were performed for AP and CP, controlling for current age, sex, and self-reported race. The reported odds ratios are relative to type O blood. Confidence intervals are 95% confidence intervals.
